# Consensus Guideline for the Management of Patients with Peritoneal Mesothelioma

**DOI:** 10.1101/2024.04.10.24305432

**Authors:** PSM Writing Group, PSM Consortium Group, Kiran K Turaga

## Abstract

**Background:** Treatment of peritoneal mesothelioma (PeM) poses significant challenges owing to its rare incidence, heterogeneity and limited clinical evidence. This manuscript describes results from a national consensus aimed at addressing management of PeM.

**Methods:** An update of the 2018 Chicago Consensus Guidelines was conducted using a Modified Delphi technique, encompassing two rounds of voting. The levels of agreement for various pathway blocks were assessed.

**Results:** Of 101 participants responding in the first round of Modified Delphi voting, 95 (94%) responded in the second round. Over 90% consensus was achieved in 5/6 and 6/6 pathway blocks in rounds I and II, respectively. Observation was recommended for benign neoplasms, with guidance for interventions in the presence of symptoms or concerning clinicopathologic features. For malignant pathology, management was outlined based on a multidisciplinary assessment of patient characteristics, disease histology, and predictive success of medical and surgical interventions. Additional emphasis was placed on multimodal therapy for Intermediate-Risk and appropriate High-Risk patients. A rapid review demonstrated limited availability of data and inconclusive findings regarding optimal systemic therapy timing. There was unanimous support for considering clinical trial enrollment.

**Conclusions:** Given limited evidence, the consensus-driven pathway provides essential guidance regarding the management of PeM. To further direct clinical care, additional dedicated research to generate higher-quality evidence is needed.

## INTRODUCTION

Peritoneal mesothelioma (PeM) represents a spectrum of neoplasms arising from epithelial and mesenchymal cells of the peritoneum^1^. PeM is remarkably rare, with an overall incidence in the United States of 0.1/100,000 people, or 300-400 new diagnoses per year^2, 3^. Asbestos and other carcinogen exposures are associated with PeM development^1, 3^. Diagnosis is overall challenging, owing to nonspecific symptoms at presentation, limitations of cross-sectional imaging in characterizing peritoneal surfaces, and the need for expert pathology review^4–7^. Despite clinical advancements, PeM remains an aggressive disease with a median overall survival less than 12 months if left untreated^8–10^. At present, there remains a paucity of literature and high-level evidence to direct clinical management of PeM.

Herein, we present the results of a national consensus on the clinical management of peritoneal mesothelioma patients, updated in line with recent evidence and expert opinion. A rapid review was conducted to investigate the role and timing of systemic therapies for PeM patients. Additionally, the consensus sought to incorporate an international perspective to assess variability in practice worldwide.

## METHODS

This initiative was part of a national multidisciplinary consortium group process aimed at streamlining guidelines for the care of patients with peritoneal surface malignancies (PSM). Full consensus and rapid review methodology has been described in detail in a separate manuscript (Submitted)^11^. The major components are summarized below:

### Consensus Group Structure

The Peritoneal Mesothelioma Working Group consisted of eight experts in surgical oncology, medical oncology, and pathology (HA, MK, BL, HK, JH, AH, GN, MZ, KR). Two core group members (LB, SW) coordinated this effort. A team of three medical students and surgical trainees conducted the rapid review. Reviewer conflicts were evaluated by a surgical oncology fellow.

### Modified Delphi Process

A modified Delphi method with two rounds of voting was employed to gather feedback regarding the clinical management pathway following preliminary synthesis of major updates since the last guideline iteration. Experts rated their agreement levels on a five-point Likert scale via a Qualtrics questionnaire. A 75% consensus threshold was set, with blocks below 90% agreement undergoing further review. Simultaneously, a summary table outlining first-line systemic therapies was generated based on histology designation.

### Rapid Review of the Literature

A MEDLINE search via PubMed between January 2000 and August 2023 addressed the key question: In patients with PeM undergoing cytoreductive surgery (CRS), what are the optimal sequences and regimens of systemic therapy? A search strategy was developed and reviewed by a medical librarian specialist, and the review protocol was pre-registered in PROSPERO (PROSPERO 2023 CRD42024519208). The search strategy included review of PubMed studies performed on humans from any date until 06/15/2023. The search strategy included the following keywords: “peritoneal mesothelioma” OR “mesothelioma” OR “Mesothelioma, Malignant” AND “surgery peritoneal” OR “surgery peritoneum” OR “resection peritoneal” OR “resection peritoneum” OR “Cytoreduc” OR “CRS” OR “intraperitoneal chemotherapy” OR “HIPEC” OR “intraperitoneal chemotherapy” OR “hyperthermic intraperitoneal chemotherapy”. Full details of the search strategy can be found in Supplemental Table 1. The Covidence platform facilitated title and abstract screening, full-text review, data extraction, and quality assessment using the Newcastle Ottawa Scale for non-randomized studies^12–14^. The review was conducted in alignment with recommendations from the Cochrane Rapid Review Methods Groups and reported in line with the Preferred Reporting Items for Systematic reviews and Meta-Analyses (PRISMA) 2020 guidelines^15, 16^.

### External Perspectives

Patient advocates within the Mesothelioma Applied Research Foundation reviewed the treatment pathway to offer patient-focused insights regarding clinical trial enrollment, research outcomes, and available resources for patients with peritoneal mesothelioma. Additionally, members of the Peritoneal Surface Oncology Group International (PSOGI) Executive Council were invited to appraise the second version of the pathway. Their comments were consolidated to evaluate alignment with global practices regarding the management of peritoneal mesothelioma.

## RESULTS

### Delphi Consensus and Rapid Review

In all, 101 field experts and thought leaders voted on the clinical pathway for PeM in the first round of the Modified Delphi method, of which 95 (94%) voted in the second round. The participants included 75 (74%) surgical oncologists, 13 (13%) medical oncologists, 10 (10%) pathologists, and 3 (3%) experts in other domains. Given the low quality of existing evidence in the literature, recommendations were based largely on expert opinion. The clinical pathway was divided into six blocks designated by key clinical aspects of management (Figure 1). The final pathway, as designated by Figure 1, represents consensus modifications to the initial proposed pathway through the Modified Delphi method (Supplemental Figure 1). Overall consensus remained high with an average of 92.8% and 97.5% of participants in agreement for Round 1 and 2, respectively (Tables 1-2).

**Figure 1:**
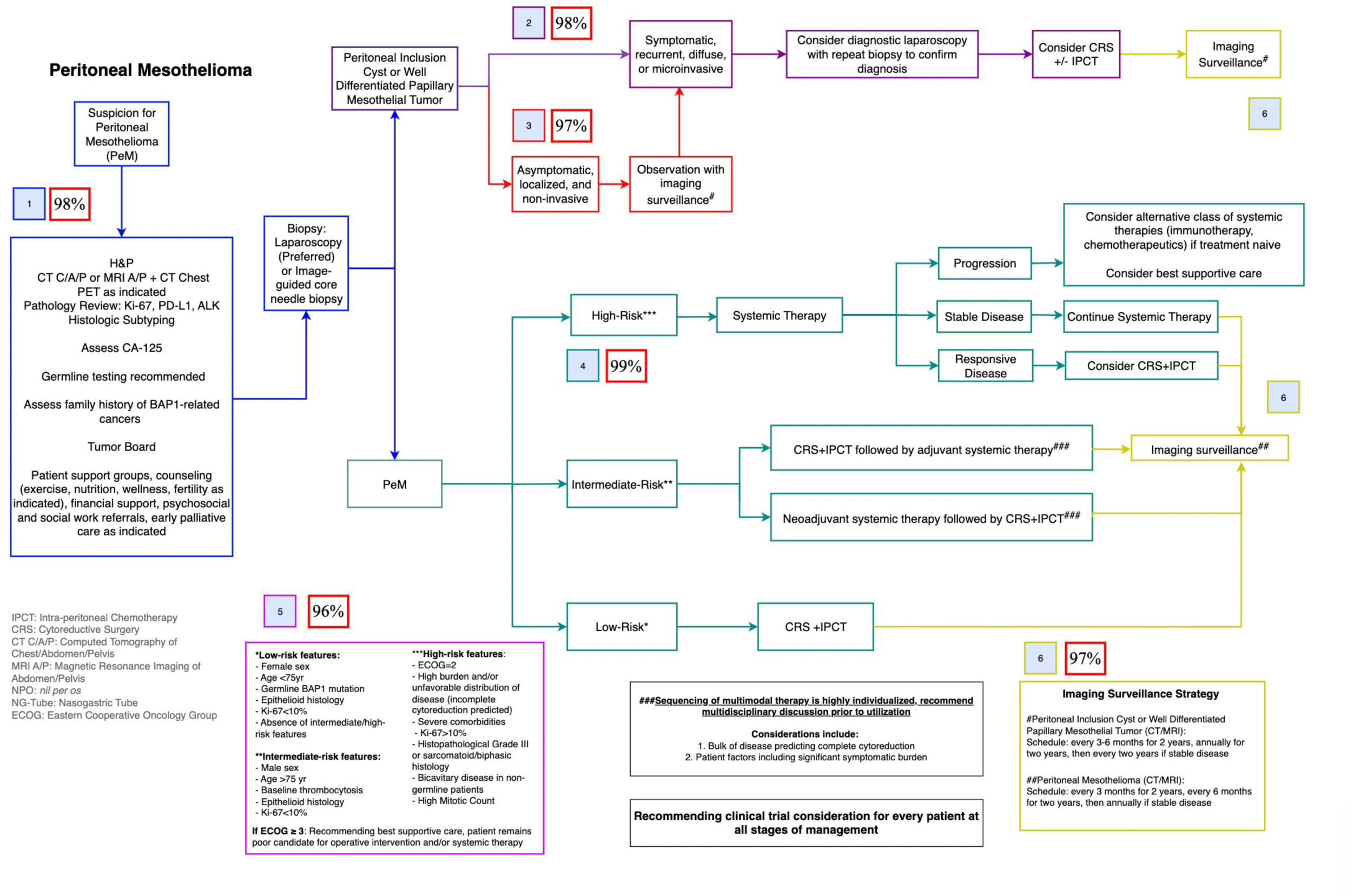
Pathway for the management of peritoneal mesothelioma.

**Table 1:** Agreement tables for the two rounds of the Modified Delphi consensus. Percentage agreement corresponds to the proportion of responses marked as ‘Strongly Agree’ or ‘Agree’ per block.

|  | **Strongly Agree** | **Agree** | **Neither Agree nor Disagree** | **Disagree** | **Strongly disagree** | **Total** | **% Agree** |
| --- | --- | --- | --- | --- | --- | --- | --- |
| **Block 1** | 67 | 29 | 2 | 3 | 0 | 101 | 95% |
| **Block 2** | 62 | 33 | 5 | 1 | 0 | 101 | 94% |
| **Block 3** | 62 | 34 | 5 | 0 | 0 | 101 | 95% |
| **Block 4** | 56 | 36 | 8 | 1 | 0 | 101 | 91% |
| **Block 5** | 54 | 35 | 11 | 1 | 0 | 101 | 88% |
| **Block 6** | 62 | 33 | 6 | 0 | 0 | 101 | 94% |

**Table 2:** Agreement tables for the two rounds of the Modified Delphi consensus. Percentage agreement corresponds to the proportion of responses marked as ‘Strongly Agree’ or ‘Agree’ per block.

|  | **Strongly Agree** | **Agree** | **Neither Agree nor Disagree** | **Disagree** | **Strongly disagree** | **Total** | **% Agree** |
| --- | --- | --- | --- | --- | --- | --- | --- |
| **Block 1** | 88 | 5 | 1 | 1 | 0 | 95 | 98% |
| **Block 2** | 85 | 8 | 1 | 0 | 1 | 95 | 98% |
| **Block 3** | 88 | 4 | 2 | 0 | 1 | 95 | 97% |
| **Block 4** | 86 | 8 | 1 | 0 | 0 | 95 | 99% |
| **Block 5** | 79 | 12 | 2 | 2 | 0 | 95 | 96% |
| **Block 6** | 86 | 6 | 3 | 0 | 0 | 95 | 97% |

A rapid review assessing the role and timing of systemic therapy in PeM revealed 588 abstracts, of which 29 full texts were reviewed, and 11 studies were included for data extraction and quality assessment (Figure 2 – PRISMA Diagram). The included studies are summarized in Table 3 and incorporated into Block 4. Summary of updates and results from the Modified Delphi consensus and rapid review are detailed below.

**Figure 2:**
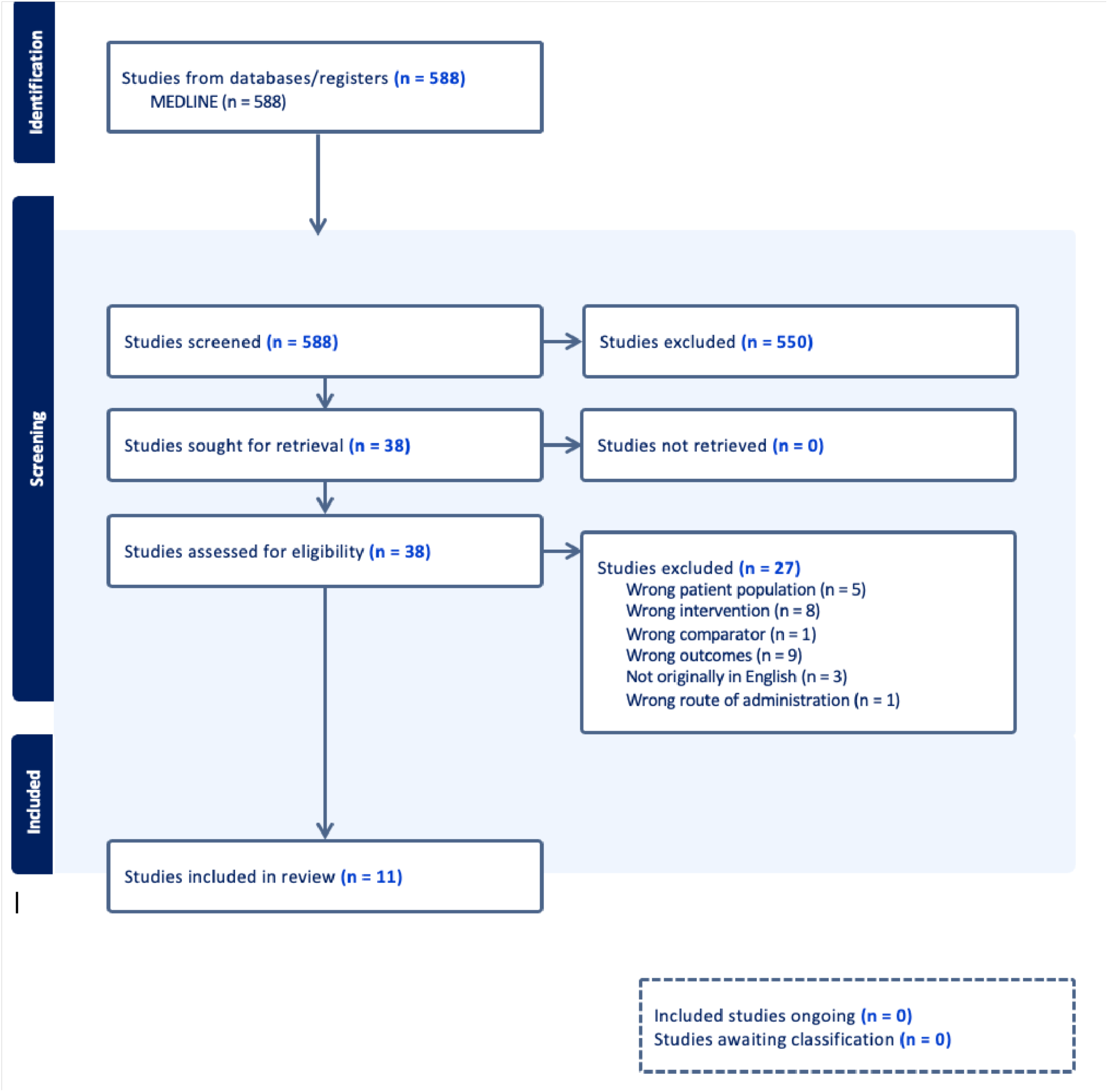
PRISMA flow diagram for the rapid review investigating the optimal sequences and regimens of systemic therapy in peritoneal mesothelioma patients undergoing CRS.

**Table 3:**
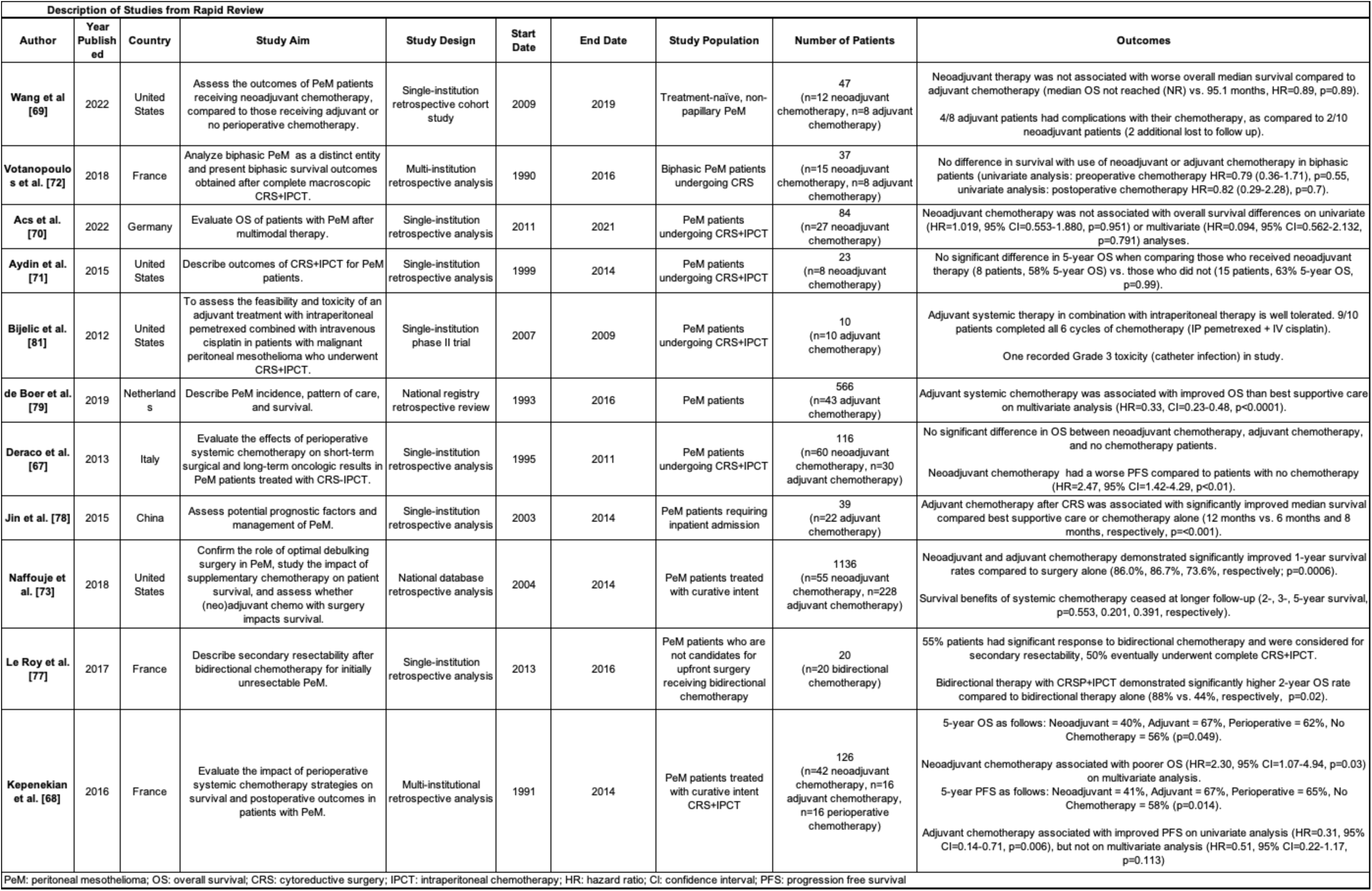
Summary of included studies within rapid review investigating role and timing of systemic therapies in PeM patients undergoing CRS.

### Summary of Major Updates

Building upon the 2018 Chicago Consensus Guidelines, the current approach involves a more stringent consensus and review methodology while engaging a larger spectrum of experts and patient advocates^17^. Major updates emphasize the importance of a comprehensive preoperative evaluation and the role of a multidisciplinary team for diagnosis, management, and surveillance of PeM patients. In addition, the new pathway also incorporates patient-focused, psychosocial interventions in preparation for possible medical and surgical intervention.

Management is outlined based on underlying histology, incorporating additional patient and disease-related factors. Observation is recommended for benign pathologies, including Peritoneal Inclusion Cysts and Well Differentiated Papillary Mesothelial Tumors (WDPMT); additional recommendations for concerning patient and tumor features are now reported. Surgical resection is reserved for patients who are sufficiently symptomatic or for those with recurrent, diffuse, or microinvasive disease. Repeat tissue diagnosis is recommended to confirm pathology, as these features are concerning for underlying malignancy. For malignant variants, the current guideline incorporates risk stratification based on underlying patient factors and predictive success of medical and surgical interventions in addition to histologic subtypes. More granular recommendations are made for Intermediate- and High-Risk categories, which are additional subgroup recommendations based on disease factors defined to further direct care. Imaging-based surveillance strategies are now defined for each tumor histology.

Given the limited randomized controlled data for treatment of PeM, the updated guideline emphasizes the importance of ongoing research efforts. Clinical trial enrollment should be considered at every step of treatment, not only to expand our knowledge of this rare disease but also to enhance patient care opportunities.

### Block 1: Diagnostic and Preoperative Considerations

#### (Agreement: Round 1 – 95%, Round 2 – 98%)

Comprehensive preoperative evaluation includes a thorough history and physical examination, diagnostic workup, and multidisciplinary tumor board discussion. Diagnostic imaging of choice includes CT or MRI of the abdomen/pelvis with oral and intravenous contrast to both optimize disease detection and radiographic approximation of tumor burden, as diffuse intra-abdominal spread portrays a risk for significant visceral organ involvement^18–20^. Although extra-abdominal metastasis is rare, CT Chest is recommended to evaluate for bicavitary disease^9, 21^. PET scans may be considered on an individual patient basis; however, the utility of this adjunct imaging has not been associated with improved outcomes^9, 22, 23^.

Diagnosis requires tissue confirmation to assess depth of invasion for proper pathological review^9, 24^. Laparoscopic approach is preferred over image-guided core needle biopsy, as the extent of intra-abdominal dissemination can be approximated and aids assessment of surgical candidacy for cytoreduction. Cytology is discouraged, as this method is unlikely to provide enough material for review and does not maintain tumor architecture^25–27^. Pathological review remains difficult and should be performed by an expert pathologist to ensure accurate diagnosis^7, 28^. Histological subtype and nuclear grade have been shown to be independent predictors for patient outcomes, therefore these determinations are imperative for proper patient assessment^9, 29, 30^. In addition, immunohistochemistry (IHC) for Ki-67, programmed death ligand-1 (PD-L1), and anaplastic lymphoma kinas (ALK) translocation is recommended, as these markers have also been associated with patient outcomes and may influence treatment recommendations. High Ki-67 (>9%) indices are associated with increased tumor invasion, along with worse median survival^30–32^. High PD-L1 expression is associated with more aggressive histologies (sarcomatoid, biphasic) but prognostic importance appears subtype specific as epithelioid tumors with high expression demonstrate improved survival^33, 34^. Currently, it remains unclear if PD-L1 status impacts candidacy for immune checkpoint inhibitors (ICI). A 2018 phase II trial investigating the utility of pembrolizumab in previously treated PeM demonstrated PD-L1 staining positively correlated with progression free survival (PFS)^35^. However, in an expanded cohort, pembrolizumab demonstrated similar efficacy in both PD-L1-positive and -negative patients, and therefore merits additional study^36^. ALK translocations represent a potential genomic aberration to exploit for treatment applicable to peritoneal—but not pleural—mesothelioma. According to a 2018 report, ALK positive PeM was found in approximately 3% of patient genomes and was associated with female sex and younger age at diagnosis^37^.

For initial evaluation, patients are recommended to undergo additional serologic and molecular testing for comprehensive assessment. Elevated CA-125 is present in approximately 50% of PeM patients and is associated with epithelioid histology and increased tumor burden^38, 39^. Germline testing is recommended for every patient, including assessment of BRCA1 associated protein (*BAP1*) mutations, as this familial syndrome has been detected in up to 25% of PeM patients and conveys an increased propensity of developing certain cancers including PeM^40–43^. At present, tumor somatic sequencing is not routinely integrated into diagnostic assessment. Recent genomic landscape studies of PeM have identified other frequently mutated genes, such as *NF2*, *SETD2*, *TP53*, thought to underlie pathogenesis; however, additional research is needed to clarify if these aberrations are clinically-useful biomarkers^44, 45^.

Additionally, establishment of a comprehensive patient support network is highly encouraged. This includes social work referrals and early palliative care as indicated. Formal evaluation by a multidisciplinary team or Tumor Board is critical to guide appropriate steps in management.

### Block 2: Benign Histology Management: Symptomatic, Progressive, Diffuse, or Microinvasive Disease

#### (Agreement: Round 1 – 94%, Round 2 – 98%)

Benign or borderline lesions, including peritoneal inclusion cysts and WDPMTs, are rare and remain poorly understood. The etiology of peritoneal inclusion cysts is currently debated, as many of these multiloculated cysts are found in young female patients with a previous abdominal surgical history, suggesting that these lesions may be reactive rather than neoplastic^46, 47^. Presenting signs and symptoms are typically related to mass effect, including palpable abdominal mass and distension^48^. Although indeterminate, peritoneal inclusion cysts are believed to have borderline to low malignant potential, with only a few reported cases of transformation in the literature^49^. WDPMT are typically multifocal lesions also found in young females^50^. WDPMT pathogenesis is unknown, and as a result, it is unclear if this lesion represents a true benign neoplasm or a precursor lesion for epithelioid PeM^51, 52^. However, recent investigations have demonstrated that WDPMT and PeM are molecularly distinct, with different genomic profiles and IHC staining, including retained BAP1 nuclear expression in WDPMT^53–55^. Despite our limited understanding, these benign lesions are noted to be phenotypically indolent compared to PeM with the majority of cases found incidentally^56^.

Diagnosis of benign lesions requires careful review by an expert pathologist to ensure no missed malignant disease. Importantly, mesothelioma in-situ (MIS), a premalignant lesion, histologically resembles WDPMT; therefore, accurate diagnosis remains critical for patient management^55, 57^. MIS is genetically similar to PeM, including *BAP1* and methylthioadenosine phosphorylase (*MTAP*) mutations, and thought to progress to invasive disease^58–60^. At present, there is no evidence to guide management of MIS.

In contrast to PeM, peritoneal inclusion cysts and WDPMT are indolent and therefore are amenable to observation with frequent outpatient follow up^47, 61, 62^. For patients with benign lesions that are sufficiently symptomatic or demonstrate evidence of progressive, diffuse, or microinvasive disease, additional evaluation is required, due to the ongoing concern for underlying malignant histology. Repeat histological examination is recommended to confirm diagnosis^55^. Patients may also be considered for cytoreductive surgery (CRS) with or without intraperitoneal chemotherapy (IPCT) after multidisciplinary re-evaluation and discussion with the patient. The role of IPCT in benign disease is traditionally thought to prevent possible malignant transformation and potentially decrease risk of local recurrence; however, its utility remains controversial^61, 63, 64^. Of note, large symptomatic peritoneal inclusion cysts are highly recurrent and surgical intervention may provoke additional lesions^47^. Systemic chemotherapy is not routinely implemented in patients with benign lesions. In one study, *Lee et al.* demonstrated that WDPMT patients receiving adjuvant therapy did not have evidence of recurrence; however, this study was limited by sample size^65^. Given the indolence of disease, this consensus does not recommend systemic therapy for peritoneal inclusion cysts or WDPMT. Patients with benign histology should undergo imaging surveillance (as designated in Block 6).

### Block 3: Benign Histology Management: Asymptomatic, Localized, Non-Invasive Disease

#### (Agreement: Round 1 – 95%, Round 2 – 97%)

Patients with demonstrated benign histology are more often asymptomatic and have localized, noninvasive disease. As benign histology is not historically associated with a decrease in survival, observation is recommended^62^. Imaging surveillance dictated by underlying histology is recommended and outlined in Block 6. If a patient has evidence of significantly progressive or microinvasive disease or becomes sufficiently symptomatic, the patient should undergo clinical re-evaluation, as designated in Block 2. Progressive, invasive, or symptomatic disease raises the concern for the presence of undetected PeM^62^.

### Block 4: PeM Management by Risk Stratification

#### (Agreement: Round 1 – 91%, Round 2 – 99%)

The term ‘mesothelioma’ implies malignant variants, therefore previously used designations of ‘diffuse’ and/or ‘malignant’ are not necessary when referring to PeM. In contrast to benign pathologies, PeM is associated with significant mortality and disease aggressiveness is highly dependent on the underlying histology as mentioned in the 2018 Chicago Consensus Guidelines^66^. In addition to histologic subtypes, the current pathway stratifies patient risk and subsequent management based on patient and disease factors and the anticipated adequacy of resection. Risk stratification should be determined by a multidisciplinary group of surgical and medical oncologists.

Currently, there is no consensus on timing or utility of systemic therapy in PeM patients. Prior studies are largely non-randomized, single institution retrospective analyses notable for significant selection bias due to the rarity and heterogeneity of PeM subtypes. To address this deficit, a rapid review was implemented to investigate the available literature. Our review of 11 appropriate studies indicated a significant paucity of available and applicable data for analysis. Therefore, meta-analysis was unsuitable due to significant heterogeneity between studies. A descriptive summary of aggregated studies and quality assessment are included (Table 3, Table 4, respectively). Additional recommendations regarding systemic therapy are provided below.

**Table 4:**
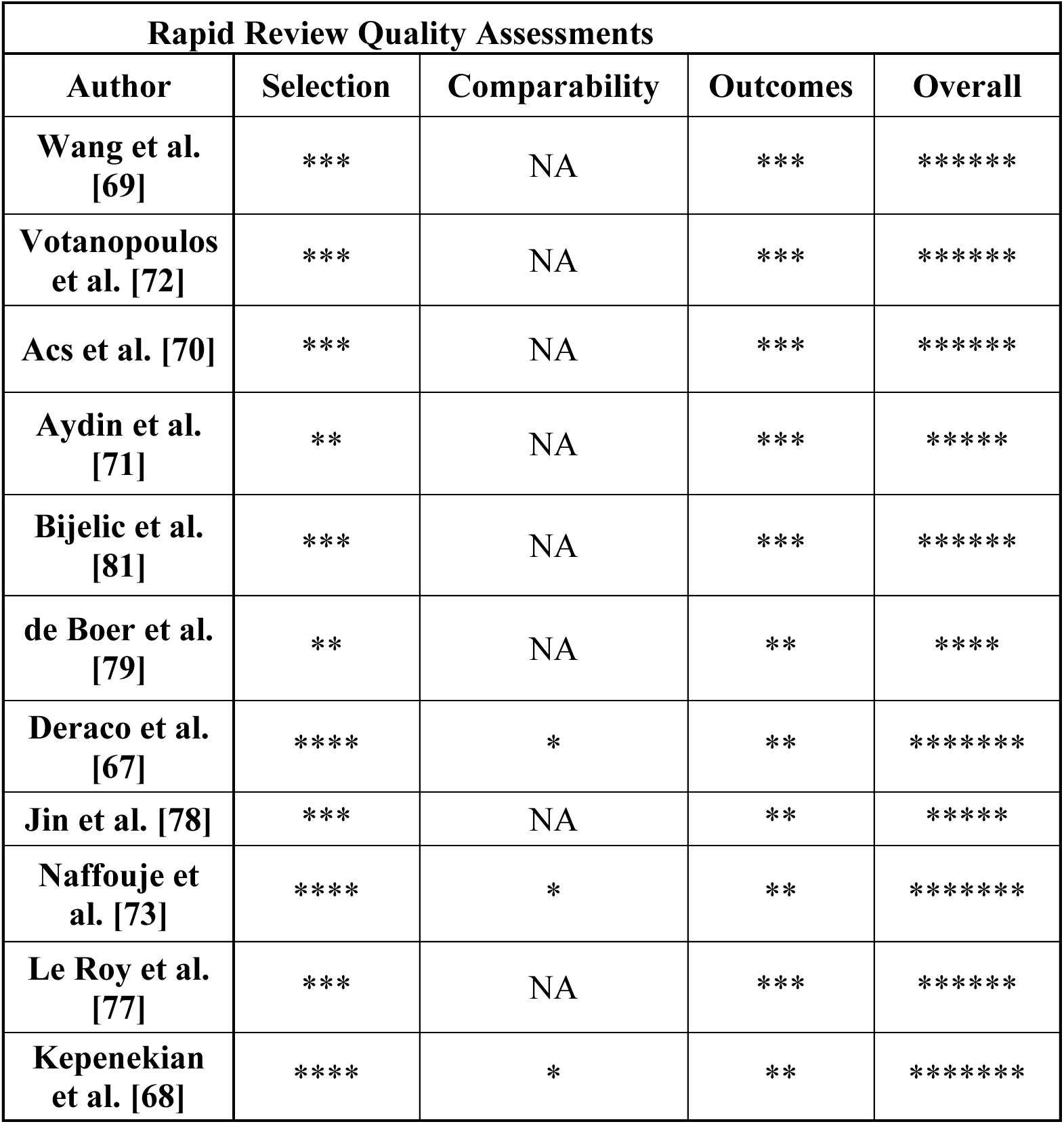
Quality assessments for included studies using the Newcastle Ottawa Scale for Cohort Studies.

Low-Risk patients are those with resectable disease and minimal perioperative risk. Current literature remains mixed for the utility of neoadjuvant therapy. In two studies, *Deraco et al.* and *Kepenekian et al.,* patients who received neoadjuvant systemic therapy demonstrated significantly worse progression free survival (PFS) and overall survival (OS), potentially attributable to selection bias due to aggressive disease biology in the neoadjuvant groups^67, 68^. However, several other studies noted no survival difference, decrement or improvement, with neoadjuvant therapy^69–72^. One study, *Naffouje et al.* demonstrated improved 1-year survival with neoadjuvant therapy; however, this OS benefit was nullified upon additional follow-up at longer time intervals^73^. Therefore, as surgical resection is strongly associated with prolonged survival, Low-Risk patients should proceed to upfront surgical intervention with CRS+IPCT^74–76^. Details of the approach to CRS-IPCT are outlined below in “Principles of Surgery”. Adjuvant therapy may be reserved for those with concerning tumor or patient risk factors and for those with incomplete cytoreduction (Intermediate- or High-Risk patients).

Intermediate-Risk disease is best approached with multimodal therapy consisting of systemic therapy and surgical resection. Considerations for systemic therapy timing include bulk of disease predicting incomplete cytoreduction and patient factors including symptomatic burden that may influence perioperative care. The role of neoadjuvant therapy in facilitating surgical resection remains controversial. *Le Roy et al.* demonstrated an overall decreased tumor volume after neoadjuvant therapy, thus allowing previously unresectable patients to undergo complete cytoreductio*n*^77^. However, additional studies by *Deraco et al.* and *Kepenekian et al.* did not demonstrate improved completeness of cytoreduction^67, 68^. Similar to neoadjuvant therapy, adjuvant therapy demonstrated inconsistent effects on survival between studies. Adjuvant therapy was associated with significantly improved median survival (*Jin et al.*) and PFS (*Kepenekian et al.* and *de Boer et al.*); however, in *Jin et al.* and *de Boer et al.*, adjuvant treatment was compared to best supportive care alone without surgical intervention, therefore confounding the survival benefits^68, 78, 79^. Several other studies did not demonstrate an overall survival benefit with adjuvant therapy, including *Deraco et al., 2013.*^67, 72, 73^. Notably, in a recent January 2024 publication, an expanded, propensity score-matched cohort including the initial *Deraco et al.* patients demonstrated that adjuvant therapy was associated with improved OS on multivariate analysis, warranting additional study^80^. Neoadjuvant and adjuvant regimens were overall well tolerated in the included studies, with the exception of *Wang et al.* in which adjuvant systemic therapy patients exhibited more complications requiring cessation or transition of therapy as compared to those receiving neoadjuvant therapy^67–69, 81^. As the current literature remains inconsistent, patient treatment plans regarding multimodal therapy should be highly individualized after discussion with a multidisciplinary team. Additional research is warranted to further delineate the role of perioperative systemic therapy.

High-Risk patients comprise a group with significant perioperative risk, high burden of disease, or with tumors of aggressive histology that would benefit from systemic therapy alone. Choice of systemic therapy, either chemotherapy or immunotherapy, is largely directed by underlying tumor biology and patient factors, and patients should undergo formal medical oncology assessment prior to initiation of therapy. Details of systemic therapy strategies are outlined below in “Principles of Systemic Therapy”. Disease response and patient quality of life after initiation of systemic therapy should be closely monitored. For High-Risk patients with evidence of disease progression while on systemic therapy, transition to an alternative therapy should be considered, along with goals of care discussion. If significant response to systemic therapy is demonstrated, select patients can further be considered for CRS+IPCT; however, this requires assessment by a multidisciplinary team.

### Block 5: Patient Risk Stratification Designation

#### (Agreement: Round 1 – 88%, Round 2 – 96%)

Criteria for patient risk stratification represents multidisciplinary consensus on established disease and patient factors. Underlying histology represents a major delineation of disease and disparate patient outcomes and therefore is a core component. Epithelioid histology, associated with a more favorable prognosis, is found in an estimated 75-90% of all patients^1, 82^. Sarcomatoid and biphasic variants have high recurrence rates after surgical intervention and median overall survival rates of approximately 7 and 10 months, respectively^74, 83–85^.

Demographics of Low-Risk patients include female sex and age younger than 75 years old, as these patient factors are historically associated with more favorable prognoses^4, 75, 79, 86, 87^. Germline *BAP1* mutations confer a less aggressive disease phenotype with a 7-fold improvement in 5-year survival compared to wild-type patients^88^. Low-Risk tumors demonstrate epithelioid histology and low proliferative index (Ki-67<10%), factors associated with improved patient survival^66, 89, 90^.

Intermediate-Risk designation comprises patients with similar histology but with patient factors associated with more aggressive disease. These patients are typically of male sex and at least 75 years old, patient prognostic factors associated with overall worse survival^4, 29, 73, 75, 87, 91^. Preoperative baseline thrombocytosis has been established as a poor prognostic marker associated with decreased overall survival^67, 78, 92^.

The High-Risk group represents patients who are not appropriate for surgical intervention. These patients have significant comorbidities, limited functional status (ECOG=2), aggressive histology (biphasic/sarcomatoid), or a burden of disease not amenable for resection. Of note, patients with bicavitary disease generally pursue nonoperative management; however, resection may be considered for very select patients (i.e. germline *BAP1*-mutated patients)^93–95^.

Patients with poor functional status (ECOG ≥3) are poor candidates for surgical intervention or systemic therapy; therefore, best supportive care is recommended.

### Block 6: Imaging Surveillance

#### (Agreement: Round 1 – 94%, Round 2 – 97%)

All patients are recommended to undergo imaging-based surveillance in addition to regular outpatient follow up. Surveillance strategies are dictated by underlying histology. For benign lesions, cross-sectional imaging (either CT or MRI of the abdomen and pelvis) is recommended every six months for the first two years after diagnosis. More frequent imaging (i.e. every three to six months) can be considered due to the risk of malignant transformation or missed disease^96, 97^. If disease remains stable, the interval for imaging may be extended annually for the following two years, and then every two years if unchanged. PeM requires more frequent imaging surveillance due to the increased risk of disease progression. After surgical intervention or initiation of medical therapy, all PeM patients are recommended to undergo imaging every three months for the first two years, every six months for the next two years, and finally annually if stable disease. If noted progression or concerning features, prompt comprehensive re-evaluation is required.

##### Principles of Systemic Therapy

Currently, multimodal treatment with CRS+IPCT and systemic therapy is recommended for Intermediate-Risk patients, whereas systemic therapies are the mainstay treatment for High-Risk patients. Low-Risk patients should proceed with upfront surgical resection without adjuvant therapy. In addition to patient candidacy, choice of systemic therapy is largely based on underlying tumor histology (Table 5) and clinical trial enrollment should be strongly considered for all PeM patients if available. Given the paucity of data on systemic therapy in PeM, most of the available information on its activity is extrapolated from studies performed in pleural mesothelioma. For epithelioid tumors, first-line chemotherapy includes pemetrexed and platinum therapy (cisplatin or carboplatin) with consideration of bevacizumab in patients without contraindication^98, 99^. Recent investigations such as the CheckMate743 trial have demonstrated a prolonged median survival associated with the ICIs, nivolumab and ipilimumab, in pleural mesothelioma patients^100^. Real-world evidence of this combination in PeM shows comparable efficacy^101^. For biphasic or sarcomatoid tumors, ICI have demonstrated significantly improved overall survival rates compared to chemotherapy regimens in pleural mesothelioma, therefore these agents are preferred as first-line agents for these histologies^100^. Of note, alternative regimens such as gemcitabine/platinum, pemetrexed alone, or vinorelbine regimens may be considered in select patient populations^98, 102–106^. Duration of therapy is defined by the regimen utilized. For patients responsive to pemetrexed/platinum therapy, a total of 4-6 cycles are recommended prior to transition to surveillance; these regimens may then be resumed if there is evidence of recurrent or progressive disease. ICI regimens may be utilized for a maximum of two years. Maintenance therapy in mesothelioma has not been associated with survival benefit^107^.

**Table 5:** Recommended systemic therapy regimens for PeM.

| Histology | Preferred First Line Therapy | Other First Line Therapy (Select Patient Populations) | Preferred Subsequent Line Therapy (Dependent on Initial First Line Therapy Utilized) | Adjunct Subsequent Therapy (Select Patient Populations) |
| --- | --- | --- | --- | --- |
| **Peritoneal Inclusion Cyst or WDPMT** | No Systemic Therapy Recommended |  |  |  |
| **Epithelioid** | Platinum* + Pemetrexed +/- Bevacizumab<br>Nivolumab + Ipilimumab<br>--<br>Clinical trial enrollment should be considered as applicable<sup>†</sup> | Gemcitabine/Platinum*<br>Pemetrexed<br>Vinorelbine | If ICI first-line: Platinum* + Pemetrexed +/- Bevacizumab<br>If chemotherapy first-line: Nivolumab +/- Ipilimumab | Atezolizumab/Bevacizumab<br>Gemcitabine<br>Vinorelbine |
| **Biphasic or Sarcomatoid** | Nivolumab + Ipilimumab<br>--<br>Clinical trial enrollment should be considered as applicable<sup>†</sup> | Platinum* + Pemetrexed +/- Bevacizumab<br>Gemcitabine/Platinum*<br>Pemetrexed<br>Vinorelbine | If ICI first-line: Platinum* + Pemetrexed +/- Bevacizumab<br>If chemotherapy first-line: Nivolumab + Ipilimumab | Atezolizumab/Bevacizumab<br>Gemcitabine<br>Vinorelbine |
| *Typically cisplatin, though carboplatin may be used in patients who cannot receive cisplatin<br>#Strongly consider neoadjuvant systemic therapy given high-risk feature<br><sup>†</sup>Consider clinical trial enrollment for every therapeutic stage for all PeM patients<br>Abbreviations: WDPMT: Well Differentiated Papillary Mesothelial Tumor, ICI: immune checkpoint inhibitor |  |  |  |  |

After initiation of therapy, disease response should be closely monitored. Progressive disease on active first-line therapy should prompt consideration of alternative agents for disease control. Second-line regimens, consisting of a different class of therapeutics, is recommended for patients who can tolerate subsequent therapy^108^. A combination regimen consisting of pemetrexed and platinum therapy, with consideration of bevacizumab in appropriate patients, is appropriate in patients who underwent ICI treatment as first line therapy, and ICI as subsequent therapy for patients who initially underwent pemetrexed/platinum treatment. Atezolizumab plus bevacizumab, one of the few regimens evaluated prospectively in a phase 2 trial in peritoneal mesothelioma patients, can also be considered^109^. Gemcitabine, gemcitabine plus ramucirumab, or oral or intravenous vinorelbine can be considered in select patient populations^106, 110–112^. For patients with stable or responsive disease on first- or second-line therapies, consideration for surgical resection after comprehensive review from a multidisciplinary team is appropriate.

In addition to the above regimens, chemo-immunotherapy approaches used in pleural mesothelioma may provide additional treatment options for PeM. Durvalumab, an anti-PD-L1 ICI, in combination with pemetrexed/platinum regimens, was well tolerated and associated with promising clinical outcomes in the DREAM and PrE0505 phase II trials^113, 114^. In the randomized phase 3 trial IND227, pembrolizumab in combination with cisplatin/pemetrexed demonstrated an improved median OS in unresectable pleural mesothelioma patients when compared to standard therapy^115^. Although not yet approved for PeM, chemo-immunotherapy regimens are currently being evaluated in clinical trials in this disease.

##### Principles of Surgery

For benign lesions, surgery is reserved for patients with concerning signs or symptoms as designated in Block 2. PeM warrants aggressive surgical intervention if amenable, as outlined in Block 4. The survival benefit of CRS+IPCT in PeM patients has long been established by several large-scale studies, as demonstrated in a meta-analysis by Helm et al. ^74–76^. A recent retrospective review of a large cohort of 2,683 PeM patients further demonstrated a profound survival benefit for CRS+IPCT, in which median survival was 70.1 months for patients who underwent surgical intervention compared to 3.0 months with no treatment. When stratified by histology, similar findings were observed for epithelioid histology with a median survival of 89.1 months for patients undergoing CRS+IPCT compared to 3.9 months for untreated patients^116^. The goal of CRS is complete resection (CC-0) or near complete cytoreduction (CC-1) of gross disease, otherwise intervention is unlikely to have a substantial survival benefit^74^. CRS typically requires extensive peritonectomies, as disease may be diffusely disseminated^9^. Resection of visceral organs, such as small bowel, spleen, gallbladder, and colon, may be necessary and associated patient morbidity should be considered prior to intervention. At present, there is no standard regimen for IPCT; however, platinum-based agents alone or in combination with doxorubicin or mitomycin have demonstrated survival benefits^68, 76^. The role of palliative debulking is largely undefined but may provide symptomatic relief. Any palliative intervention should be reviewed on an individual patient basis by a multidisciplinary team^24, 117^.

## DISCUSSION

Management of peritoneal mesothelioma remains clinically complex due to the rarity and heterogeneity of disease. The 2018 Chicago Consensus Guidelines instituted vital pathways for clinical management; however, additional consideration for patient and disease factors from a multidisciplinary perspective is required. By engagement of surgical oncology, medical oncology, radiology, and pathology field experts, we have established a comprehensive clinical pathway for disease management of peritoneal mesothelioma. Most notably, we emphasize the lack of high-quality evidence to direct this guideline, necessitating additional investigations and clinical trial enrollment. Despite the low level of evidence, this consensus offers valuable guidance regarding diagnosis and management of benign entities and PeM.

Major limitations of this expert consensus merit discussion. Firstly, the available evidence for our rapid review was of low quality and scarce. Therefore, the consensus methodology was employed to provide guidance regarding matters of equipoise. Secondly, the expert panel consisted primarily of surgical oncologists. Having expected this bias from the inception phases, thought leaders in medical oncology and other disciplines were involved early for reviewing feedback from the first Delphi round and outlining principles of systemic therapy. Lastly, the Delphi consensus entailed voting on blocks rather than individual itemized recommendations, aligning with the original Chicago Consensus framework. While this approach helped mitigate survey fatigue, it may have compromised the granularity of feedback received.

### PSOGI: International Perspective

Recently, the Peritoneal Surface Oncology Group International (PSOGI) issued consensus guidelines for management for peritoneal mesothelioma, peritoneal inclusion cyst, and WDPMTs ^7, 118^. Overall, this consensus is in line with established international recommendations for disease management. Both guidelines recommend the same diagnostic and preoperative evaluation for new patients; however, the guideline presented here offer additional patient support considerations. Similar to our updated guideline, PSOGI guidelines instituted patient stratification based on tumor resectability and patient factors to direct PeM management. Shared emphasis of the importance of a multidisciplinary team was present. The PSOGI guidelines also described the role of other regional perfusion modalities, namely Early Postoperative IPCT and Normothermic IPCT, which were absent in our recommendations given these approaches are not employed frequently in North America.

### Patient Advocate Perspective

The two patient advocate perspectives we obtained shared many similarities in terms of their priorities, outlook on clinical trials, and sources of support and information. They stressed the importance of clinical trials given the relatively limited therapeutic options for PeM. Neither patient participated in a clinical trial, one due to ineligibility and one due to positive response to initial CRS+IPCT. Both patients noted the high morbidity of current treatment regimens, with one patient writing that the “extensive surgery for this disease can be lifesaving but is also life changing. Many of us have continuous quality of life issues that need to be addressed.” While both patients agreed that survival should remain the most important research outcome, they emphasized the importance of continuing to reduce morbidity while advancing treatment regimens. One patient wrote that early connection to other healthcare providers (ex. mental health providers, physical therapists, other specialist physicians) is integral for adequate comorbidity management, especially as the number of patients living with PeM increases. Both patients cited peer support groups as an important source of information and comfort throughout their disease course.

## CONCLUSION

In summary, we report an updated Modified Delphi consensus on management of PeM that included a multidisciplinary team of experts, including surgical/medical oncologists, radiologists, pathologists, and patient support groups. For benign pathology, observational management with frequent surveillance is recommended. PeM necessitates CRS+IPCT for amenable disease (Low- and Intermediate-Risk patients); however, implementation of multimodal therapy or systemic therapy alone requires detailed, multidisciplinary assessment for those patients at increased perioperative risk and/or with aggressive disease (Intermediate- and High-Risk patients). Additional research is needed to advance our understanding of both disease and appropriate treatment for PeM patients.

## Data Availability

The datasets used and/or analyzed during the current study are available from the corresponding author on reasonable request.

