## Supplementary figures and images for "Consensus Guideline for the Management of Patients with Peritoneal Mesothelioma"

### Supplemental Figure 1

#### Supplemental Figure 1

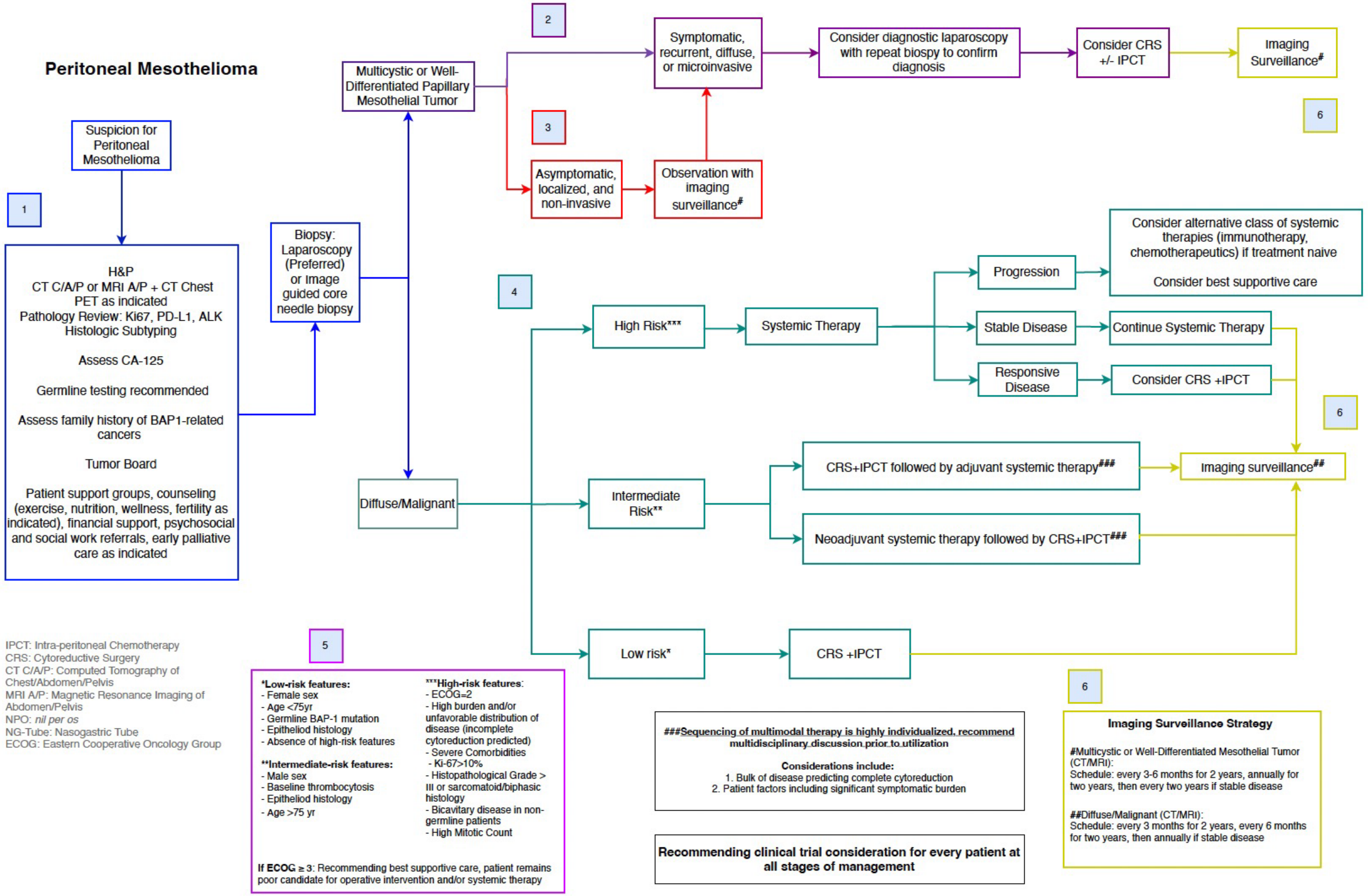
