## Supplemental Table 1 for "Consensus Guideline for the Management of Patients with Peritoneal Mesothelioma"

**SUPPLEMENTARY MATERIAL**

**Supplemental Table 1. Search Strategy for Key Question:** In patients with PeM undergoing cytoreductive surgery (CRS), what are the optimal sequences and regimens of systemic therapy?

PubMed; restrictions: any date - 2023/06/15, performed on humans

| Search line | Search term |
|---|---|
| 1 | peritoneal mesothelioma[tw] OR mesothelioma[MeSH Major Topic] OR "Mesothelioma, Malignant"[MeSH Major Topic] |
| 2 | ("surgery peritoneal"[tiab:~5]) OR ("surgery peritoneum"[tiab:~5]) OR ("resection peritoneal"[tiab:~5]) OR ("resection peritoneum"[tiab:~5]) OR (Cytoreduc*[tw] OR CRS[tw]) OR intraperitoneal chemotherapy*[tw] OR HIPEC*[tw] OR intraperitoneal chemotherapy*[tw] OR (hyperthermic intraperitoneal chemotherapy[Mesh Major Topic]) |
| 3 | #1 AND #2 |
